# Association of Seizure with COVID-19 Vaccines in Persons with Epilepsy: A Systematic Review and Meta-analysis

**DOI:** 10.1101/2023.05.06.23289604

**Authors:** Ali Rafati, Melika Jameie, Mobina Amanollahi, Mana Jameie, Yeganeh Pasebani, Delaram Sakhaei, Saba Ilkhani, Sina Rashedi, Mohammad Yazdan Pasebani, Mohammadreza Azadi, Mehran Rahimlou, Churl-Su Kwon

**Author notes:** **Corresponding author:** Churl-Su Kwon M.D., M.P.H., FRSPH., 622 West 168th Street, PH19-106, New York, NY 10032, (T), (E). These two authors contributed equally (Ali Rafati, Melika Jameie).

## Abstract

**Objective:** Seizure following immunization, especially in persons with epilepsy (PwE), has long been a concern, and seizure aggravation followed by Coronavirus Disease 2019 (COVID-19) vaccines is a serious issue for PwE. The immunization rate in PwE has been lower compared to same-age controls due to vaccine hesitancy and concerns about seizure control. Herein, we systematically reviewed the seizure activity-related events in PwE following COVID-19 vaccination.

**Methods:** Four search engines were searched from inception until January 31, 2023, and the Preferred Reporting Items for Systematic Reviews and Meta-Analyses was followed. Random- and fixed-effect models using the logit transformation method were used for meta-analysis. The quality of the studies was evaluated by the Newcastle-Ottawa scale. Outcomes of interest included (a) pooled proportion of increased seizure frequency and (b) pooled incidence proportion of status epilepticus (SE) in PwE receiving COVID-19 vaccines.

**Results:** Of the 2207 studies identified, 18 met eligibility criteria, of which 16 entered the meta-analysis. The pooled proportion of increased seizure frequency (16 studies-4197 PwE) was 5% (95CI: 3%-6%, I^2^ =57%), further subcategorized into viral vector (3%, 95CI: 2%-7%, I^2^ =0%), mRNA (5%, 95CI: 4%-7%, I^2^ =48%), and inactivated (4%, 95CI: 2%-8%, I^2^ =77%) vaccines. The pooled incidence proportion of SE (15 studies-2480 PwE) was 0.08% (95CI: 0.02%-0.32%, I^2^ =0%), further subcategorized into the viral vector (0.00%, 95CI: 0.00%-1.00%, I^2^ =0%), mRNA (0.09%, 95CI: 0.01%-0.62%, I^2^ =0%), and inactivated (0.00%, 95CI: 0.00%-1.00%, I^2^ =0%) vaccines. No significant difference was observed between mRNA and viral vector vaccines (5 studies, 1122 vs. 198 PwE, respectively) regarding increased seizure frequency (OR: 1.10, 95CI: 0.49-2.50, p-value=0.81, I^2^ =0%).

**Significance:** The meta-analysis proposed a 5% increased seizure frequency following COVID-19 vaccination in PwE, with no difference between mRNA and viral vector vaccines. Furthermore, we found a 0.08% incidence proportion for SE. While this safety evidence is noteworthy, this cost should be weighed against vaccination benefits.

## 1. Introduction

Coronavirus disease 2019 (COVID-19) is a highly transmissible disease caused by the severe acute respiratory syndrome coronavirus 2 (SARS-CoV-2), which was first reported on December 31, 2019 ^1^. The World Health Organization declared COVID-19 a pandemic on March 11, 2020 ^2^. Nine months later, in December 2020, the U.S. Food and Drug Administration (FDA) approved two vaccines for COVID-19 protection ^3^. Early evaluations of vaccine effectiveness were promising, with lower transmission rates, hospitalization, and death among vaccinated individuals ^4^. No serious side effects were reported during the vaccines’ phase 3 clinical trials, with chills, headache, injection site pain, fatigue, and myalgia being the most commonly reported side effects ^3^. Currently, except for severe allergic reactions to the previous vaccine dose or to a component of the COVID-19 vaccine, there are no contraindications for COVID-19 vaccine administration ^5^.

Current recommendations encourage specific populations, such as immunocompromised people, pregnant/lactating women, and people with a history of other allergies, to get the vaccine after a risk assessment and appropriate counseling ^6^. Recent studies have also recommended that COVID-19 vaccines are generally safe and well-tolerated in persons with epilepsy (PwE), indicating a favorable view of vaccination against COVID-19 in this population ^7^. However, seizures following immunization have long been a source of conflict; while some vaccines may increase the likelihood of seizures, getting vaccinated at the correct time can reduce seizures by preventing infections ^8^.

Some vaccines, such as the measles-mumps-rubella (MMR) ^8, 9^, the measles-mumps-rubella-varicella (MMRV) combination ^8^, diphtheria-tetanus toxoids-whole-cell pertussis (DTwP) ^9^, and diphtheria-tetanus toxoids-acellular pertussis-inactivated poliovirus-Haemophilus influenza type b (DTaP-IPV-Hib) ^10^ may increase the risk of seizure, mainly by causing fever. This concern is more prominent in PwE, with numerous research focusing on the increased risk of post-vaccination seizures in this group ^11, 12^. Albeit the concerns, PwE are generally advised to be vaccinated against vaccine-preventable diseases, such as the flu, because the risk of seizures and status epilepticus (SE) increases during the infection ^13^.

As with other vaccines, seizure aggravation following COVID-19 vaccination is a serious issue for PwE. Concerns about losing seizure control have resulted in increased vaccine hesitancy and reduced vaccine administration rate in PwE compared to their same-age controls ^14^. Herein, we systematically reviewed the literature on seizure-related activities (i.e., increased seizure frequency, change in seizure type, and SE incidence) after receiving COVID-19 vaccination in PwE. We also conducted meta-analyses to calculate the proportion of post-vaccination increased seizure frequency and SE incidence.

## 2. Methods

### 2.1. Objectives

We systematically reviewed post-COVID-19-vaccination seizure activity-related events reported in the literature, including increased seizure frequency, change in seizure type, and SE occurrence. Increased seizure frequency and SE incidence were also investigated by pooling effect estimates as follows: (1) studies reporting increased seizure frequency following the COVID-19 vaccines; (2) studies reporting SE incidence following the COVID-19 vaccines; and (3) studies comparing mRNA vs. viral-vector COVID-19 vaccines in terms of post-vaccinal increased seizure frequency.

### 2.2. Study design and search strategy

This study was carried out in accordance with the guidelines of the Preferred Reporting Items for Systematic Reviews and Meta-Analyses (PRISMA) ^15^. The study protocol was registered with PROSPERO (CRD42022312475). A systematic search was performed in MEDLINE (via PubMed), Web of Science, Scopus, Cochrane Library, and Google Scholar from the inception of COVID-19 (December 2019) to January 31, 2023. To identify studies not found through the traditional searching/screening process, we also searched review publications, editorials, letters to editors, and conference papers, along with the references of the included studies. The main search terms included “SARS-CoV-2 vaccine”, “COVID-19 vaccine”, “epilepsy”, “seizure”, and “convulsion” (Supplementary Table S1). No restriction was made based on the age of the participants or the literature language. As the previously published data were used, the study was exempt from ethical approval by the local institutional ethics board.

### 2.3. Eligibility criteria

The study participants were PwE, having undergone COVID-19 vaccination with any COVID-19 vaccine platform, including mRNA, viral vector, inactivated virus, protein subunit, etc. Studies reporting < 10 PwE receiving COVID-19 vaccines were excluded. No restriction was made based on the age of the participants or the literature language. The outcomes of interest were any seizure-related adverse events following COVID-19 vaccination (increased seizure frequency, SE incidence, seizure type change, etc.). The studies were included only if the seizure diagnosis was confirmed by a neurologist/healthcare professional or fulfilled the clinical criteria of the International League Against Epilepsy ^16^ and/or the International Classification of Diseases (9^th^ or 10^th^ revision), as mentioned by each study.

### 2.4. Selection process

The obtained records were first transferred to the EndNote. After duplicate removal, two independent researchers (MeJ and YP) screened all articles. Firstly, all references were screened independently by their titles and abstracts. Secondly, two reviewers independently screened the full-text articles of abstracts identified in the first phase. Finally, the records not meeting the preestablished eligibility criteria were excluded. Any disagreement regarding the inclusion of articles was resolved by the consensus of a third party as required.

### 2.5. Data extraction

The following data of interest were extracted: (a) study-specific variables, including the first author’s name, publication year, sample size, and study design, (b) vaccine-related variables, including vaccine type, number of received doses, and the interval between vaccination and outcomes of interest, (c) demographic, clinical, and epilepsy-related variables including age, sex, history of COVID-19 infection, epilepsy duration, epilepsy type, baseline seizure frequency, and anti-seizure medications (ASMs), and (d) the number of patients presenting with post-COVID-19-vaccination seizure activity-related events.

### 2.6. Quality assessment

The quality of the studies was assessed using the Newcastle-Ottawa scale (NOS) for cross-sectional studies and cohorts^17^. Studies with a score of ≥6 were considered high quality. Two independent researchers (DS and MA) conducted the quality assessment, and conflicts were resolved via consensus.

### 2.7. Data synthesis

The quantitative synthesis was performed in three sections. The first section pooled the studies reporting increased seizure frequency after COVID-19 vaccination, subcategorized by vaccine platforms. The second section compared the increased seizure frequency between mRNA and viral-vector vaccines. In the third section, the SE incidence after COVID-19 vaccines was pooled and further subcategorized by vaccine platforms.

### 2.8. Statistical analysis

All statistical analyses were performed with the R package ‘meta’ version 5.2-0, R version 4.2.1 ^18^, using the Logit transformation method (single-arm proportions) and Mantel-Haenszel method ^19, 20^ (for dichotomous data). To pool the single-arm proportions and for dichotomous data on seizure incidence, the ‘metaprop’ and ‘metabin’ functions were used, respectively. The incidence proportion and odds ratio (OR), with the 95% confidence intervals (CIs), were calculated for the measure of effect.

The between-study heterogeneity was evaluated by Cochran’s Q statistic, τ2 using the restricted maximum-likelihood (REML) estimator and I^2^ index ^21, 22^. It was decided to choose between the fixed-effects or random-effects models based on the I^2^ index. If I^2^≥50%, hence substantial heterogeneity, the random-effects model was used; otherwise, the fixed-effects model was used. Publication bias was assessed with funnel plots, and asymmetry was tested using Egger’s and Peter’s tests ^23, 24^. A two-tailed p-value<0.05 was considered significant.

## 3. Results

### 3.1. Study selection and characteristics

Of the 2207 records obtained from the systematic search after duplicate removal, 18 studies were included in the qualitative synthesis (PRISMA flow diagram in Figure S1). Of the 18 studies, nine were cohort ^25-33^, and nine were cross-sectional ^14, 34-41^ and were conducted in China ^14, 26, 29, 30, 32-34, 37^, Italy ^25^, the United Kingdom ^41^, the United States ^35, 40^, Turkey ^36^, Germany ^39^, Spain ^27^, Kuwait ^38^, Mexico ^28^, and Australia ^31^. The vaccine platforms utilized included mRNA (Pfizer/BioNTech and Moderna), viral-vector (Janssen, Oxford/AstraZeneca, and CanSino), inactivated virus (CoronaVac and Sinopharm), and adjuvanted protein subunit (Zifivax) vaccines.

Table 1 summarizes the studies’ characteristics, including demographic data, vaccine platform, and main findings on seizure-activity-related events. Other clinical and epilepsy-related characteristics, including the history of COVID-19 infection, epilepsy duration, age of epilepsy onset, epilepsy type, baseline seizure frequency, and baseline ASMs.

**Table 1.**
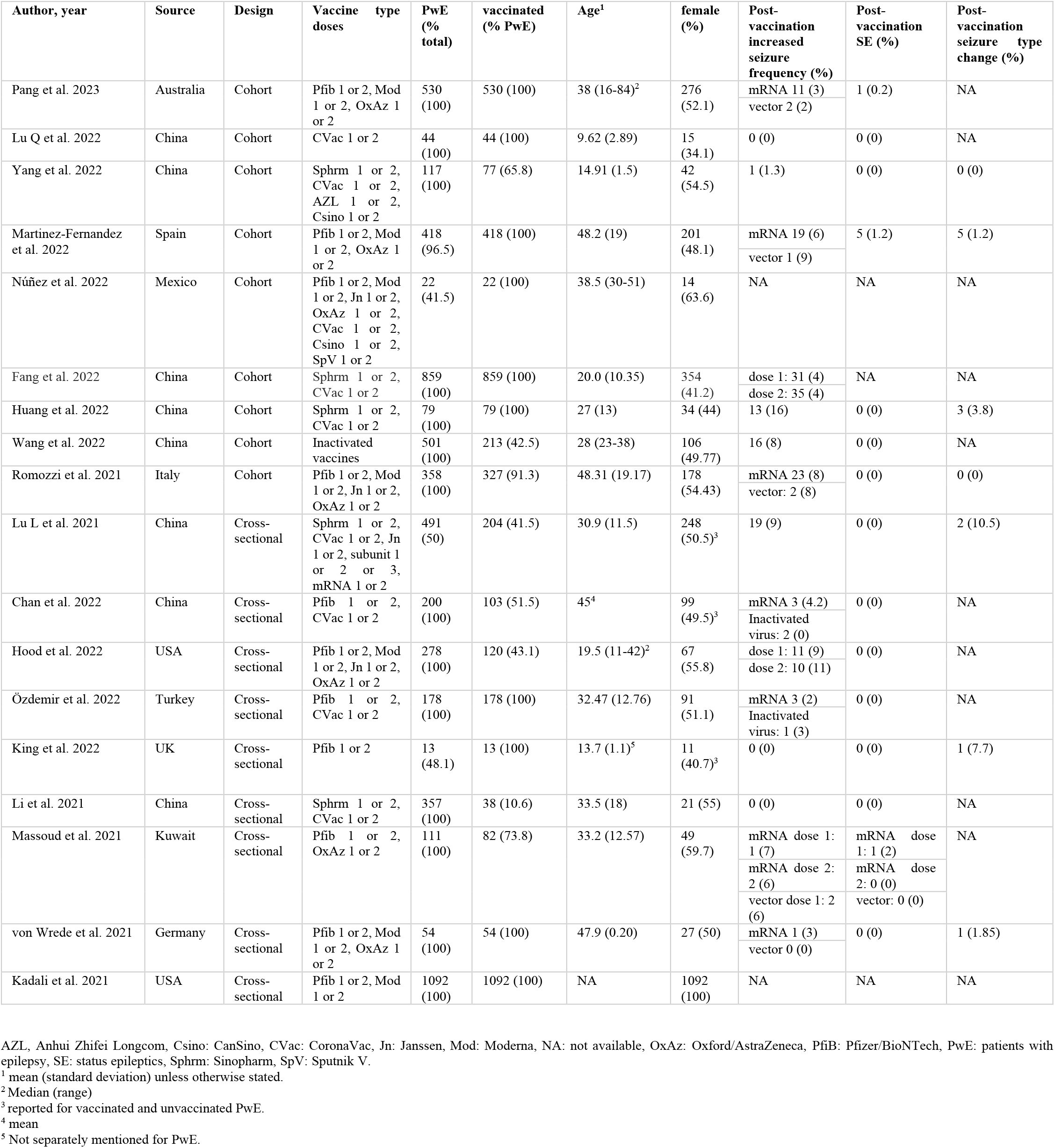
Summary of the studies.

### 3.2. Quality appraisal

In Supplementary Tables S2 and S3, the quality assessment of the included studies is reviewed. All records showed NOS scores of ≥6 except for one ^41^.

### 3.3. Increased seizure frequency

Sixteen studies reporting increased seizure frequency were pooled in this section ^14, 25-27, 29-39, 41^. The meta-analysis was performed on three subgroups of vaccine platforms (Figure 1). The pooled increased seizure frequency per PwE was 3% (95CI: 2%-7%, I^2^=0%; Cochran Q p-value=0.68) for viral-vector (6 studies ^25, 27, 31, 36, 38, 39^, 233 PwE), 5% (95CI: 4%-7%, I^2^=48%; Cochran Q p-value=0.04) for mRNA (9 studies ^25, 27, 31, 34-36, 38, 39, 41^, 1556 PwE), and 4% (95CI: 2%-8%, I^2^=77%; Cochran Q p-value<0.01) for the inactivated virus (8 studies ^14, 26, 29, 30, 32-34, 37^, 2408 PwE) vaccines.

**Figure 1.**
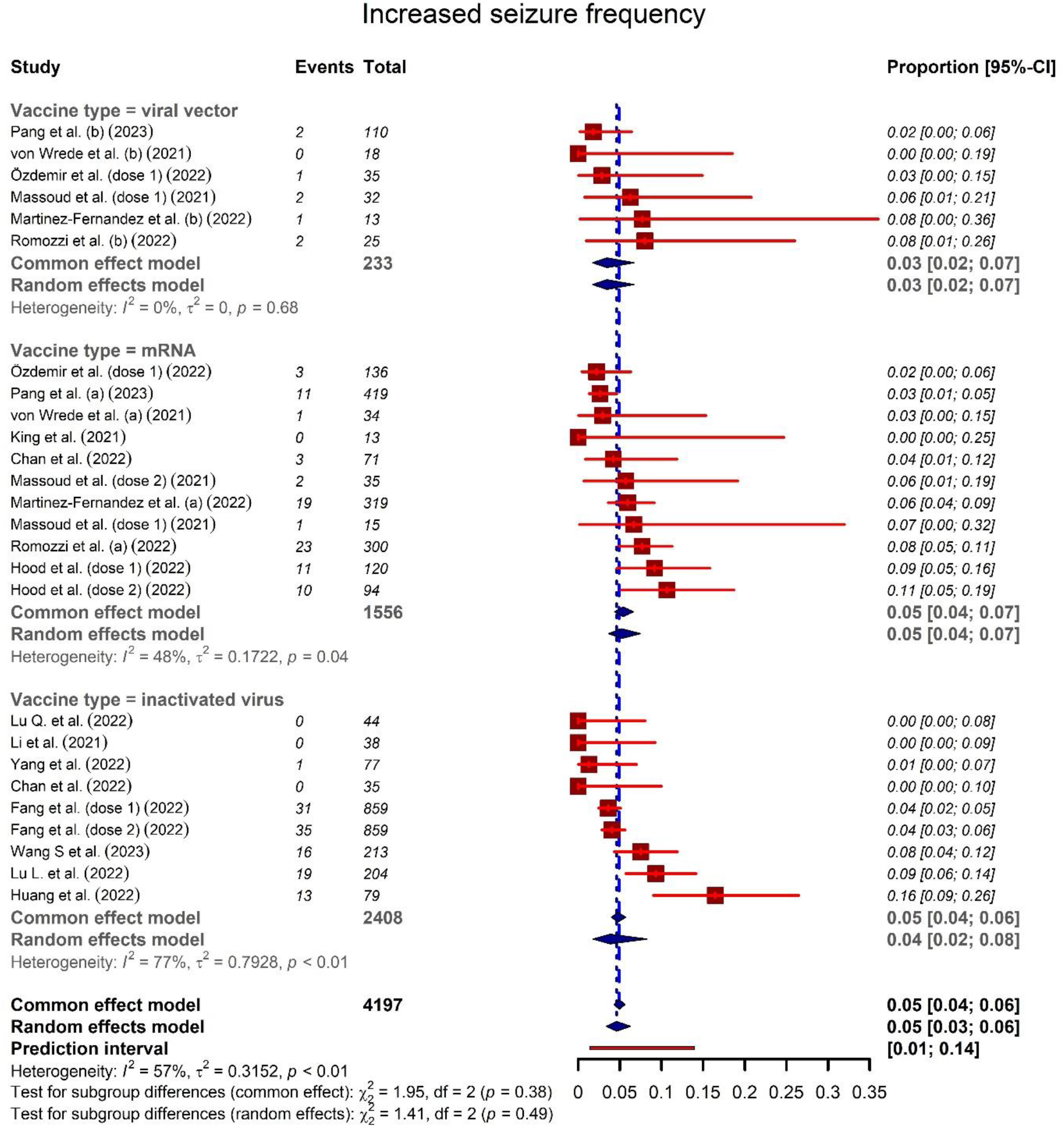
Forest plot representing the proportion of increased seizure frequency after COVID-19 vaccines.

All vaccines combined (16 studies ^14, 25-27, 29-39, 41^, 4197 PwE), the pooled increased seizure frequency was calculated at 5% (95CI: 3%-6%, I^2^=57%; Cochran Q p-value<0.01). Overall, no publication bias was observed (funnel plot in Supplementary Figure S2) with Egger’s and Peter’s tests’ p-value of 0.34 and 0.27, respectively.

### 3.4. Increased seizure frequency: mRNA vaccines vs. viral-vector vaccines

Five studies reported the post-vaccination data on both mRNA and viral-vector vaccines, separately (Figure 2) ^25, 27, 31, 38, 39^. In this meta-analysis, the odds of increased seizure frequency between 1122 mRNA-vaccine-receiving PwE and 198 viral-vector-vaccine-receiving PwE were not significantly different (OR: 1.10, 95CI: 0.49-2.50, p-value= 0.81; I^2^=0%; Cochran Q p-value=0.99). No publication bias was detected with Egger’s and Peter’s tests’ p-values of 0.89 and 0.96, respectively (funnel plot in Supplementary Figure S3).

**Figure 2.**
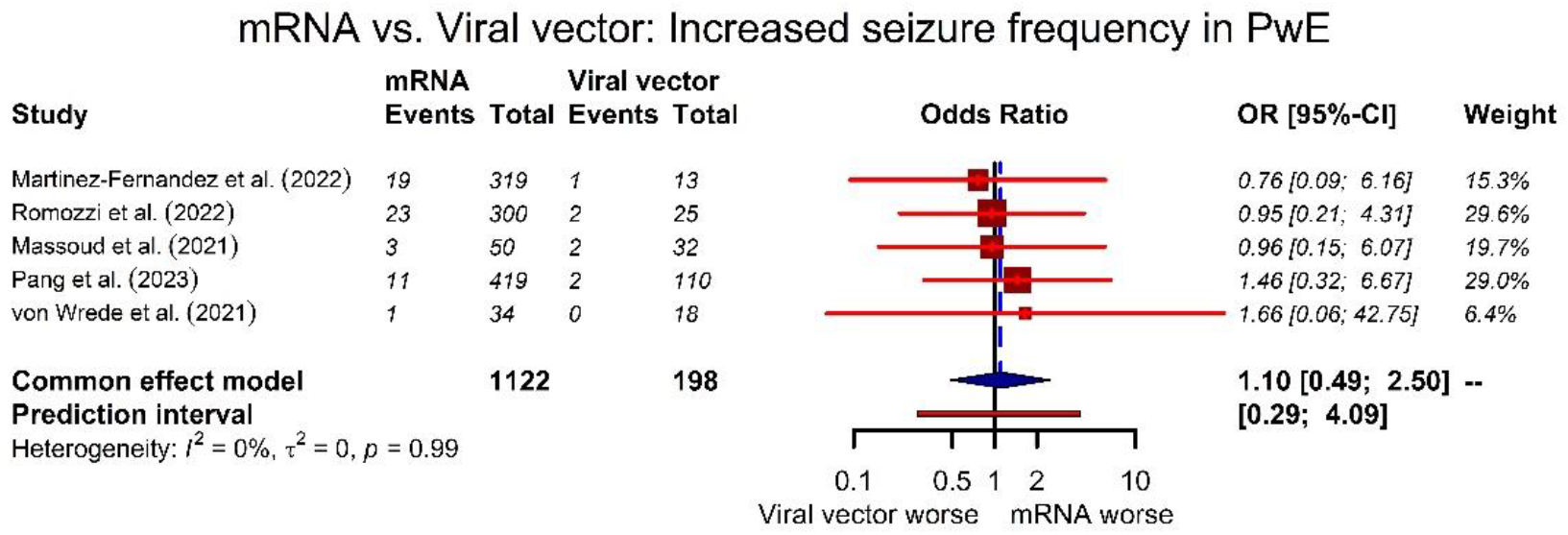
Forest plot representing the comparison between mRNA vs. viral-vector COVID-19 vaccines in terms of increased seizure frequency.

### 3.5. SE frequency

SE was reported in 15 studies ^14, 25-27, 30-39, 41^. The meta-analyses were carried out on three subgroups of vaccine platforms (Figure 3). The pooled SE incidence proportion per PwE was 0.09% (95CI: 0.01%-0.62%, I^2^=0%; Cochran Q p-value =1.00) for mRNA (8 studies^25, 27, 34-36, 38, 39, 41^, 1137 PwE), 0.00% (95CI: 0.00%-1.00%, I^2^=0%; Cochran Q p-value =1.00) for the inactivated virus (7 studies ^14, 26, 30, 32-34, 37^, 690 PwE), and 0.00% (95CI: 0.00%-1.00%, I^2^=0%; Cochran Q p-value=1.00) for viral vector vaccines (5 studies ^25, 27, 36, 38, 39^, 123 PwE). All vaccine platforms combined (15 studies ^14, 25-27, 30-39, 41^, 2480 PwE), the pooled SE incidence proportion per PwE was 0.08% (95CI: 0.02%-0.32%, I^2^=0%; Cochran Q p-value=1.00). No publication bias was observed with Egger’s test p-value of 0.53 (funnel plot in Supplementary Figure S4).

**Figure 3.**
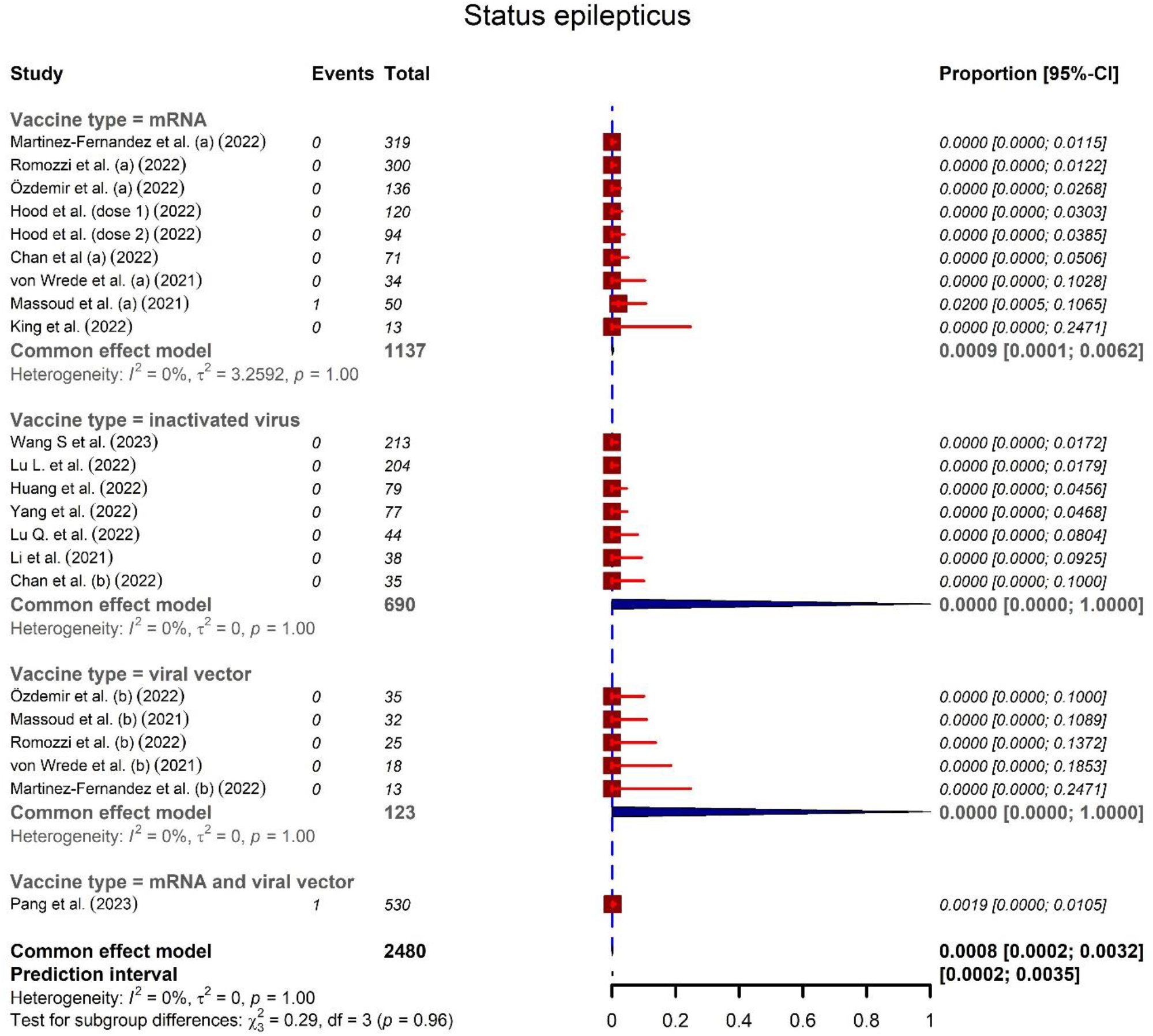
Forest plot representing pooled status epilepticus incidence proportion after COVID-19 vaccines.

## 4. Discussion

According to our meta-analysis of 16 studies, accounting for 4197 PwE, the pooled proportion of increased seizure frequency following COVID-19 vaccination in PwE was 5%. The subgroup analysis showed an increased seizure frequency of 3% after the viral vector, 5% after the mRNA, and 4% after the inactivated virus vaccines in PwE. Following COVID-19 vaccination, the pooled SE incidence proportion from 15 studies, accounting for 1480 PwE, was 0.08%. According to the subgroup analysis, the SE incidence proportion was 0.00% after the viral vector, 0.09% after the mRNA, and 0.00% after the inactivated virus vaccines. No significant difference was detected between the viral vector and mRNA vaccines in terms of increased seizure frequency.

Although the exact etiology behind developing seizures after COVID-19 vaccination remains unclear, fever ^34-36^, ASM-related factors (non-adherence, not using bridge ASMs, number of ASMs, change in ASM type or dose, etc.) ^14, 25^-^27^, ^32, 33, 35, 39^, pre-vaccine baseline seizure status ^25^, ^27, 31, 33^, genetic factors ^34^, immune-related factors ^35^, hyponatremia ^42^, metabolic disturbances ^27, 31^, infections ^27^, tumors ^27^, stress ^27^, insomnia or disturbed sleep routine ^27, 33^, fatigue ^32^, and stimulant drinks such as caffeine ^32^, have been mentioned in the literature as possible post-COVID-19 vaccination seizure predisposing factors, which could be related to vaccines or not. ASM-related factors, fever, and baseline seizure status were among the most discussed factors.

### ASM-related factors

Martinez-Fernandez *et al*. studied 418 PwE, of whom 26 patients experienced increased seizure frequency ^27^. The authors did not find any evident cause for decompensation other than vaccination in 28.5% of patients. Among the patients with another possible cause for decompensation, ASM-related factors (missed dose, change doses, malabsorption diseases) were found in most patients (38.5%) ^27^. Other suggested possible causes for decompensation were metabolic disturbances (3.8%), infections (3.8%), tumors (7.7%), and stress or insomnia (7.7%) ^27^. According to von Wrede *et al*. study on 54 adult PwE, irregular ASM intake was simultaneously reported by those who developed seizure-activity-related events (increased seizure frequency in two patients, increased seizure intensity in one, and change in seizure type in one) ^39^. Consistently, according to Lu *et al*. study on 491 PwE, about one-third of the patients reporting increased seizure frequency had reduced or stopped their ASMs due to the fear of potential interaction with vaccines ^14^. Huang *et al*. also indicated that improper ASM administration was significantly associated with seizure worsening ^33^. Univariate and logistic regression analysis in Wang *et al*. study indicated that fewer ASM numbers and poor ASM adherence were associated with post-COVID-19 vaccination seizure worsening in PwE ^32^. On the other hand, Romozzi *et al*. suggested that being treated with a higher number of ASMs was associated with seizure worsening following COVID-19 vaccination ^25^. In another study on 117 patients with benign childhood epilepsy with centrotemporal spikes, one patient developed a post-COVID-19 vaccination seizure within three months after receiving CanSino: Ad5-nCoV. According to the authors, she was not taking her medications regularly in the days before the seizures, although previously taking them regularly for over a year ^26^. However, the authors emphasized that there is still no conclusive evidence confirming any causal relationship between COVID-19 vaccination and developing seizures in children with epilepsy who are not taking their medications ^26^.

Using bridge ASMs was another factor discussed in the literature; according to the experts’ consensus, bridge ASMs are considered prophylactic measures for reducing the possibility of post-vaccination seizures, particularly in patients with Dravet syndrome (DS, also known as severe myoclonic epilepsy of infancy [SMEI]) ^43^. However, in the Hood *et al*. study, patients who reported using bridge ASM following vaccination were more likely to report increased seizure activity ^35^. The authors hypothesized that patients who had worse baseline seizure control or who were more sensitive to seizure triggers may have used preemptive prophylactic measures, including a bridge ASM, more frequently ^35^. Concisely, data regarding ASM-related factors, such as patients’ ASM adherence during the vaccination period, number of ASMs, change in ASMs or adding a new ASM, and using bridge ASMs should be taken into account when evaluating vaccine-related adverse events in PwE.

### Post-vaccination fever

Fever and other forms of hyperthermia frequently cause seizures in patients with DS ^44^. In their study on 278 patients with DS, Hood *et al*. reported that fever was identified by some caregivers as a direct trigger for seizure ^35^. However, several studies have reported that fever is not always present in patients with DS who have had vaccine-triggered seizures, suggesting the possible role of other immune-related pathways in post-vaccination seizure etiology ^35, 45^. Consistently, another study indicated that while fever was reported more often following the 2^nd^ dose of the COVID-19 vaccine (D2) than the 1^st^ dose (D1), increased seizure following the two doses was comparable (9% and 11%, respectively) ^35^. On the other hand, in the Hood *et al*. study, patients who used antipyretics, compared to those who did not, were more likely to report increased seizure activity after COVID-19 vaccination, while several caregivers reported that antipyretics helped reduce fever and, consequently, seizure activity ^35^. According to the authors, this might be indicative of antipyretics being used reactively rather than proactively, making it challenging to interpret the causal relationship ^35^. According to another study with 200 PwE who had received COVID-19 vaccination, three patients experienced seizure-related events, of whom 2 had developed fever, and one had an afebrile seizure ^34^. Based on this descriptive observation, the authors suggested that fever might be a possible risk factor for seizure worsening following COVID-19 vaccination ^34^. Consistently, in another study, all patients with post-vaccination seizures (4 of 178 PwE) had systemic adverse events, especially fever ^36^. On the other hand, in Pang *et al*. study, PwE with post-D1 seizure exacerbation and those without post-D1 seizure exacerbation had comparable rates of systemic side effects, including fever ^31^. Similarly, none of the 14 patients with post-vaccination seizure worsening in a study reported fever following COVID-19 vaccination ^33^. Summarily, the role of fever on developing seizures following COVID-19 vaccination requires further studies, considering confounding factors, such as the baseline seizure status, ASM status, sleep status, stress level, etc.

### Pre-vaccination baseline seizure status

According to one study, the only factor significantly associated with increased seizure frequency following COVID-19 vaccination was the baseline seizure frequency ^27^; while being seizure free for more than a year before vaccination showed a protective role, having monthly seizures -defined as 1-3 seizures/month-prior to vaccination was a significant risk factor for developing post-vaccination seizure ^27^. According to the authors, no other clinical variables, including the number of ASMs, epilepsy type or etiology, drug-resistant epilepsy (DRE), active epilepsy, epilepsy surgery, previous history of COVID-19, vaccine type, or the number of received doses, were related to an increase in seizure frequency ^27^. Consistently, Romozzi *et al*. study indicated an association between seizure worsening following COVID-19 vaccination and baseline seizure frequency patterns ^25^. Of note, while univariate analysis indicated an association between DRE and seizure worsening, multivariable regression did not indicate such an association in this study ^25^. In line with these studies, according to Pang *et al*., being seizure-free for 12 months at the time of the first dose was the only indicator of not having COVID-19 vaccine-associated seizures ^31^. However, it is worth noting that this study was more relevant to drug-resistant PwE, and the results might not be representative of the general population of PwE^31^.

### Genetic and immune-related factors

Among 200 patients in Chan *et al*. study, all three patients who experienced seizure worsening after COVID-19 vaccination had generalized epilepsy with genetic etiologies, suggesting that idiopathic/genetic etiology might be a risk factor for post-COVID-19 vaccination seizure worsening ^34^. On the other hand, Hood *et al*. suggested that immune-related pathways might play a role in seizure causation following vaccination ^35^, which has also been suggested by previous research on vaccines other than COVID-19 ^46^.

### Other possible related factors

According to Martinez-Fernandez *et al*. and Huang *et al*. studies, insomnia and disturbed sleep routine were associated with post-COVID-19 vaccination seizure frequency ^27, 33^. These findings were in line with previous studies that proposed a lack of sleep as an independent triggering factor for developing seizures ^47-49^. Having stimulant drinks, such as common coffee, after COVID-19 vaccination was another seizure precipitant in PwE, suggested by Wang *et al*. study ^32^. In this sense, previous evidence supports the effect of caffeine intake on seizure control in PwE ^50^. Stress and fatigue were other seizure precipitants following COVID-19 vaccination, suggested by Wang *et al*. and Martinez-Fernandez *et al*. studies, respectively ^27, 32^. These results corroborate previous research regarding seizure precipitants ^51^. Metabolic and electrolyte disturbances were other suggested triggering factors for seizure worsening following COVID-19-vaccination ^31, 42^. Among 13 patients who developed post-D1 seizure activity in Pang *et al*. study, one patient with type 1 diabetes mellitus experienced vomiting and diarrhea that resulted in metabolic abnormalities and hypoglycemia, possibly resulting in their post-D1 seizure ^31^. A case report suggested deep hyponatremia as another possible rare trigger for seizures following COVID-19 vaccination ^42^. Ten days after receiving the Pfizer/BioNTech vaccine, the patient developed a generalized tonic-clonic seizure, which was diagnosed as neuroleptic malignant syndrome (NMS). The concurrent serum sodium level was 111 mEq/L, suggesting that hyponatremia, which may have been caused by the immunization, may have contributed to the seizure ^42^. Developing hyponatremia after Oxford/AstraZeneca COVID-19 vaccination has also rarely been reported ^52^.

### Vaccination benefits and risks

Importantly, the chance of developing seizures after COVID-19 vaccines seems to be lower than COVID-19 infection ^53^. According to a meta-analysis of case-series and retrospective cohorts, among 71 PwE, 49 patients presented with seizures as an initial manifestation of COVID-19 infection with an incidence of 69% (54%-85%, p=0.10, I^2^=34%), which is much higher than the rate we reached in the present meta-analysis ^53^. In addition, while other vaccines, other than COVID-19 vaccines, have also been shown to be possibly associated with developing seizures in PwE ^54-56^, they are advised to be vaccinated against vaccine-preventable diseases due to immunization benefits. Notably, the incidence proportion of increased seizure frequency found in this study (5%) does not suggest causality between the COVID-19 vaccine and seizures. Therefore, in the authors’ opinion, the possible minor increase in the risk of developing a seizure following immunization is outweighed by the vaccinations’ benefits.

### Limitations

The present study has several limitations. First, no randomized clinical trials were available to include in the meta-analysis. Second, since we used the published data, not the individual-patient-level data, we could not conduct subgroup analyses based on parameters such as sex, age, and vaccine dose. Third, the majority of the studies did not provide sufficient data on other risk factors related to seizure occurrence, such as drug history, medication compliance, comorbid diseases, sleep status, and metabolic disorders. Fourth, it should be noted that the recorded seizures following vaccination might be partly associated with a reporting bias resulting from the excessive concern and attention to detecting adverse events of the vaccines. Moreover, the results may be vulnerable to recall bias since most studies relied on patients’ self-reported data. Lastly, some studies did not provide an exact definition for seizure exacerbation; hence, this definition could differ among studies. The same limitation exists for the data regarding the duration between vaccination and seizure-related events. Altogether, the mentioned limitations might have developed potential confounders for the meta-analysis.

## Conclusion

In sum, available COVID-19 vaccines seem safe and appropriate for PwE in the COVID-19 era. According to our meta-analysis of to-date published studies, the overall proportion of seizure-related adverse events among PwE was not considerable. Taken together, a minor increase in seizure risk after COVID-19 vaccination cannot be ruled out; however, the benefits seem to outweigh the potential risks.

Of note, due to the correlations between vaccines, fever, and seizures in many PwE, recommending antipyretic medications might be considered a preventative strategy for seizure worsening after vaccination. Proper ASM administration and adherence to the prescribed ASMs could also reduce the rates of post-vaccination seizures in PwE. In addition, data regarding the possible interactions between epilepsy medications and COVID-19 vaccines should be provided for the patients to avoid both adverse events and unnecessary pauses in taking medications due to the fear of interactions. Patients with poorer baseline seizure control are possibly at higher risk for developing seizures following COVID-19 vaccination and should be monitored closely. Additionally, patients whose health deteriorated because of unstable cortical excitability caused by poor ASM adherence, disturbed sleep routine, fatigue, or stimulant drinks may be more prone to be affected by vaccinations. Therefore, after COVID-19 vaccination, PwE should be instructed to avoid possible seizure triggers such as sleep deprivation, stimulant drinks, fatigue, and anxiety. More thorough investigations are required to identify factors associated with epilepsy-related adverse events following COVID-19 vaccination.

## Data Availability

All data produced in the present work are contained in the manuscript

## Supplementary information

The supplementary materials are available in the supplemental file.

## Author contribution

**Ali Rafati:** conceptualization, formal analysis, investigation, data curation, methodology, visualization, wrote the original draft, reviewed, and edited the manuscript.

**Melika Jameie:** conceptualization, formal analysis, investigation, data curation, methodology, visualization, wrote the original draft, reviewed, and edited the manuscript.

**Mobina Amanollahi:** investigation, data curation, methodology, visualization, wrote the original draft, reviewed, and edited the manuscript.

**Mana Jameie:** investigation, data curation, methodology, visualization, wrote the original draft, reviewed, and edited the manuscript.

**Yeganeh Pasebani:** investigation, data curation, methodology, visualization, wrote the original draft, reviewed, and edited the manuscript.

**Delaram Sakhaei:** investigation, data curation, reviewed and edited the manuscript.

**Saba Ilkhani:** investigation, data curation, reviewed and edited the manuscript.

**Sina Rashedi:** investigation, data curation, reviewed and edited the manuscript.

**Mohammad Yazdan Pasebani:** investigation, data curation, reviewed and edited the manuscript.

**Mohammadreza Azadi:** investigation, data curation, reviewed and edited the manuscript.

**Mehran Rahimlou:** validated, reviewed, and edited the manuscript.

**Churl-Su Kwon:** project administration, supervised, validated, reviewed, and edited the manuscript.

All authors read and approved the final manuscript.

## Data availability

No original data to share. All analyses were performed on previously published articles.

## Acknowledgments

None.

