## Supplementary file for "Association of Seizure with COVID-19 Vaccines in Persons with Epilepsy: A Systematic Review and Meta-analysis"

### *Corresponding author:

[†] These two authors contributed equally.

### Table S1- Systematic search syntax for databases with results as of January 31, 2023.

| MEDLINE (via PubMed) | | |
|---|---|---|
| No. | Syntax | No. of results |
| 1. | ("COVID-19"[Title/Abstract] OR "sars cov 2"[Title/Abstract] OR "covid 19 vaccines"[Title/Abstract] OR "covid 19 vaccines"[MeSH Terms] OR "covid 19 vaccine*"[Title/Abstract] OR "sars cov 2 vaccine*"[Title/Abstract] OR "sars cov 2 vaccine*"[Title/Abstract] OR "vaccines sars cov 2"[Title/Abstract]) | 301,347 |
| 2. | "Seizures"[MeSH Terms] OR "seizure*"[Title/Abstract] OR "single seizure*"[Title/Abstract] OR "seizure single"[Title/Abstract] OR "seizures somatosensory"[Title/Abstract] OR "seizure somatosensory"[Title/Abstract] OR "somatosensory seizure*"[Title/Abstract] OR "vertiginous seizure*"[Title/Abstract] OR "seizures vestibular"[Title/Abstract] OR "vestibular seizure*"[Title/Abstract] OR "seizures visual"[Title/Abstract] OR "seizure visual"[Title/Abstract] OR "visual seizure*"[Title/Abstract] OR "nonepileptic seizure*"[Title/Abstract] OR "seizures nonepileptic"[Title/Abstract] OR "non epileptic seizure*"[Title/Abstract] OR "non epileptic seizure*"[Title/Abstract] OR "seizure non epileptic"[Title/Abstract] OR "generalized absence seizure*"[Title/Abstract] OR "absence seizures generalized"[Title/Abstract] OR "tonic clonic seizure*"[Title/Abstract] OR "seizure tonic clonic"[Title/Abstract] OR "Tonic Clonic Seizure"[Title/Abstract] OR "clonic seizures tonic"[Title/Abstract] OR "seizure tonic clonic"[Title/Abstract] OR "Tonic Clonic Seizures"[Title/Abstract] OR "seizures tonic clonic"[Title/Abstract] OR "generalized tonic clonic seizure*"[Title/Abstract] OR "generalized tonic clonic seizure*"[Title/Abstract] OR "seizure generalized tonic clonic"[Title/Abstract] OR "seizures generalized tonic clonic"[Title/Abstract] OR "tonic clonic seizures generalized"[Title/Abstract] OR "clonic seizure*"[Title/Abstract] OR "seizure clonic"[Title/Abstract] OR "seizures clonic"[Title/Abstract] OR "tonic seizure*"[Title/Abstract] OR "seizure tonic"[Title/Abstract] OR "seizures tonic"[Title/Abstract] OR "convulsions non epileptic"[Title/Abstract] OR "non epileptic convulsion*"[Title/Abstract] OR "convulsion*"[Title/Abstract] OR "atonic absence seizure*"[Title/Abstract] OR "absence seizure atonic"[Title/Abstract] OR "absence seizures atonic"[Title/Abstract] OR "seizure convulsive*"[Title/Abstract] OR "convulsive seizure*"[Title/Abstract] OR "seizures convulsive"[Title/Abstract] OR "seizures motor"[Title/Abstract] OR "motor seizure*"[Title/Abstract] OR "seizure motor"[Title/Abstract] OR "jacksonian seizure*"[Title/Abstract] OR "seizure jacksonian"[Title/Abstract] OR "seizures auditory"[Title/Abstract] OR "auditory seizure*"[Title/Abstract] OR "seizure auditory"[Title/Abstract] OR "seizures focal"[Title/Abstract] OR "focal seizure*"[Title/Abstract] OR "seizure focal"[Title/Abstract] OR "partial seizure*"[Title/Abstract] OR "seizure partial"[Title/Abstract] OR "seizures generalized"[Title/Abstract] OR "generalized seizure*"[Title/Abstract] OR "seizure generalized"[Title/Abstract] OR "gustatory seizure*"[Title/Abstract] OR "seizures olfactory"[Title/Abstract] OR "olfactory seizure*"[Title/Abstract] OR "myoclonic seizure*"[Title/Abstract] OR "seizure myoclonic"[Title/Abstract] OR "epileptic seizure*"[Title/Abstract] OR "seizure epileptic"[Title/Abstract] OR "seizures epileptic"[Title/Abstract] OR "seizures sensory"[Title/Abstract] OR "seizure sensory"[Title/Abstract] OR "sensory seizure*"[Title/Abstract] OR "absence seizure*"[Title/Abstract] OR "seizure absence"[Title/Abstract] OR "complex partial seizure*"[Title/Abstract] OR "partial seizure complex"[Title/Abstract] OR "partial seizures complex"[Title/Abstract] OR "seizure complex partial"[Title/Abstract] OR "atonic seizure*"[Title/Abstract] OR "seizure atonic"[Title/Abstract] | 171,994 |
| 3. | #1 AND #2 | 946 |

| Web of Science | | |
|---|---|---|
| No. | Syntax | No. of results |
| 1. | ((TI=COVID-19 OR AB=COVID-19) OR (TI="sars cov 2" OR AB="sars cov 2") OR (TI="covid 19 vaccines" OR AB="covid 19 vaccines") OR ALL="covid 19 vaccines" OR (TI="covid 19 vaccine*" OR AB="covid 19 vaccine*") OR (TI="sars cov 2 vaccine*" OR AB="sars cov 2 vaccine*") OR (TI="sars cov 2 vaccine*" OR AB="sars cov 2 vaccine*") OR (TI="vaccines sars cov 2" OR AB="vaccines sars cov 2")) | 85,018 |
| 2. | (ALL=Seizures OR (TI=seizure* OR AB=seizure*) OR (TI="single seizure*" OR AB="single seizure*") OR (TI="seizure single" OR AB="seizure single") OR (TI="seizures somatosensory" OR AB="seizures somatosensory") OR (TI="visual seizure*" OR AB="visual seizure*") OR (TI="nonepileptic seizure*" OR AB="nonepileptic seizure*") OR (TI="absence seizures generalized" OR AB="absence seizures generalized") OR (TI="tonic clonic seizure*" OR AB="tonic clonic seizure*") OR (TI="myoclonic seizure*" OR AB="myoclonic seizure*") OR (TI="epileptic seizure*" OR AB="epileptic seizure*") OR (TI="seizure absence" OR AB="seizure absence")) | 7,560 |
| 3. | #1 AND #2 | 748 |

| Scopus | | |
|---|---|---|
| No. | Syntax | No. of results |
| 1. | (TITLE-ABS(COVID-19) OR TITLE-ABS("sars cov 2") OR TITLE-ABS("covid 19 vaccines") OR INDEXTERMS("covid 19 vaccines") OR TITLE-ABS("covid 19 vaccine*") OR TITLE-ABS("sars cov 2 vaccine*") OR TITLE-ABS("sars cov 2 vaccine*") OR TITLE-ABS("vaccines sars cov 2")) | 396,247 |
| 2. | INDEXTERMS(Seizures) OR TITLE-ABS(seizure*) OR TITLE-ABS("single seizure*") OR TITLE-ABS("seizure single") OR TITLE-ABS("seizures somatosensory") OR TITLE-ABS("seizure somatosensory") OR TITLE-ABS("somatosensory seizure*") OR TITLE-ABS("vertiginous seizure*") OR TITLE-ABS("seizures vestibular") OR TITLE-ABS("vestibular seizure*") OR TITLE-ABS("seizures visual") OR TITLE-ABS("seizure visual") OR TITLE-ABS("visual seizure*") OR TITLE-ABS("nonepileptic seizure*") OR TITLE-ABS("seizures nonepileptic") OR TITLE-ABS("non epileptic seizure*") OR TITLE-ABS("non epileptic seizure*") OR TITLE-ABS("seizure non epileptic") OR TITLE-ABS("generalized absence seizure*") OR TITLE-ABS("absence seizures generalized") OR TITLE-ABS("tonic clonic seizure*") OR TITLE-ABS("seizure tonic clonic") OR TITLE-ABS("Tonic Clonic Seizure") OR TITLE-ABS("clonic seizures tonic") OR TITLE-ABS("seizure tonic clonic") OR TITLE-ABS("Tonic Clonic Seizures") OR TITLE-ABS("seizures tonic clonic") OR TITLE-ABS("generalized tonic clonic seizure*") OR TITLE-ABS("generalized tonic clonic seizure*") OR TITLE-ABS("seizure generalized tonic clonic") OR TITLE-ABS("seizures generalized tonic clonic") OR TITLE-ABS("tonic clonic seizures generalized") OR TITLE-ABS("clonic seizure*") OR TITLE-ABS("seizure clonic") OR TITLE-ABS("seizures clonic") OR TITLE-ABS("tonic seizure*") OR TITLE-ABS("seizure tonic") OR TITLE-ABS("seizures tonic") OR TITLE-ABS("convulsions non epileptic") OR TITLE-ABS("non epileptic convulsion*") OR TITLE-ABS(convulsion*) OR TITLE-ABS("atonic absence seizure*") OR TITLE-ABS("absence seizure atonic") OR TITLE-ABS("absence seizures atonic") OR TITLE-ABS("seizure convulsive*") OR TITLE-ABS("convulsive seizure*") OR TITLE-ABS("seizures convulsive") OR TITLE-ABS("seizures motor") OR TITLE-ABS("motor seizure*") OR TITLE-ABS("seizure motor") OR TITLE-ABS("jacksonian seizure*") OR TITLE-ABS("seizure jacksonian") OR TITLE-ABS("seizures auditory") OR TITLE-ABS("auditory seizure*") OR TITLE-ABS("seizure auditory") OR TITLE-ABS("seizures focal") OR TITLE-ABS("focal seizure*") OR TITLE-ABS("seizure focal") OR TITLE-ABS("partial seizure*") OR TITLE-ABS("seizure partial") OR TITLE-ABS("seizures generalized") OR TITLE-ABS("generalized seizure*") OR TITLE-ABS("seizure generalized") OR TITLE-ABS("gustatory seizure*") OR TITLE-ABS("seizures olfactory") OR TITLE-ABS("olfactory seizure*") OR TITLE-ABS("myoclonic seizure*") OR TITLE-ABS("seizure myoclonic") OR TITLE-ABS("epileptic seizure*") OR TITLE-ABS("seizure epileptic") OR TITLE-ABS("seizures epileptic") OR TITLE-ABS("seizures sensory") OR TITLE-ABS("seizure sensory") OR TITLE-ABS("sensory seizure*") OR TITLE-ABS("absence seizure*") OR TITLE-ABS("seizure absence") OR TITLE-ABS("complex partial seizure*") OR TITLE-ABS("partial seizure complex") OR TITLE-ABS("partial seizures complex") OR TITLE-ABS("seizure complex partial") OR TITLE-ABS("atonic seizure*") OR TITLE-ABS("seizure atonic") | 257,003 |
| 3. | #1 AND #2 | 1,742 |

| Cochrane | | |
|---|---|---|
| No. | Syntax | No. of results |
| 1. | (COVID-19:ti,ab OR "sars cov 2":ti,ab OR "covid 19 vaccines":ti,ab OR [mh "covid 19 vaccines"] OR ("covid 19" NEXT vaccine*):ti,ab OR ("sars cov 2" NEXT vaccine*):ti,ab OR ("sars cov 2" NEXT vaccine*):ti,ab OR "vaccines sars cov 2":ti,ab) | 13,121 |
| 2. | [mh Seizures] OR seizure*:ti,ab OR ("single" NEXT seizure*):ti,ab OR "seizure single":ti,ab OR "seizures somatosensory":ti,ab OR "seizure somatosensory":ti,ab OR ("somatosensory" NEXT seizure*):ti,ab OR ("vertiginous" NEXT seizure*):ti,ab OR "seizures vestibular":ti,ab OR ("vestibular" NEXT seizure*):ti,ab OR "seizures visual":ti,ab OR "seizure visual":ti,ab OR ("visual" NEXT seizure*):ti,ab OR ("nonepileptic" NEXT seizure*):ti,ab OR "seizures nonepileptic":ti,ab OR ("non epileptic" NEXT seizure*):ti,ab OR ("non epileptic" NEXT seizure*):ti,ab OR "seizure non epileptic":ti,ab OR ("generalized absence" NEXT seizure*):ti,ab OR "absence seizures generalized":ti,ab OR ("tonic clonic" NEXT seizure*):ti,ab OR "seizure tonic clonic":ti,ab OR "Tonic Clonic Seizure":ti,ab OR "clonic seizures tonic":ti,ab OR "seizure tonic clonic":ti,ab OR "Tonic Clonic Seizures":ti,ab OR "seizures tonic clonic":ti,ab OR ("generalized tonic clonic" NEXT seizure*):ti,ab OR ("generalized tonic clonic" NEXT seizure*):ti,ab OR "seizure generalized tonic clonic":ti,ab OR "seizures generalized tonic clonic":ti,ab OR "tonic clonic seizures generalized":ti,ab OR ("clonic" NEXT seizure*):ti,ab OR "seizure clonic":ti,ab OR "seizures clonic":ti,ab OR ("tonic" NEXT seizure*):ti,ab OR "seizure tonic":ti,ab OR "seizures tonic":ti,ab OR "convulsions non epileptic":ti,ab OR ("non epileptic" NEXT convulsion*):ti,ab OR convulsion*:ti,ab OR ("atonic absence" NEXT seizure*):ti,ab OR "absence seizure atonic":ti,ab OR "absence seizures atonic":ti,ab OR ("seizure" NEXT convulsive*):ti,ab OR ("convulsive" NEXT seizure*):ti,ab OR "seizures convulsive":ti,ab OR "seizures motor":ti,ab OR ("motor" NEXT seizure*):ti,ab OR "seizure motor":ti,ab OR ("jacksonian" NEXT seizure*):ti,ab OR "seizure jacksonian":ti,ab OR "seizures auditory":ti,ab OR ("auditory" NEXT seizure*):ti,ab OR "seizure auditory":ti,ab OR "seizures focal":ti,ab OR ("focal" NEXT seizure*):ti,ab OR "seizure focal":ti,ab OR ("partial" NEXT seizure*):ti,ab OR "seizure partial":ti,ab OR "seizures generalized":ti,ab OR ("generalized" NEXT seizure*):ti,ab OR "seizure generalized":ti,ab OR ("gustatory" NEXT seizure*):ti,ab OR "seizures olfactory":ti,ab OR ("olfactory" NEXT seizure*):ti,ab OR ("myoclonic" NEXT seizure*):ti,ab OR "seizure myoclonic":ti,ab OR ("epileptic" NEXT seizure*):ti,ab OR "seizure epileptic":ti,ab OR "seizures epileptic":ti,ab OR "seizures sensory":ti,ab OR "seizure sensory":ti,ab OR ("sensory" NEXT seizure*):ti,ab OR ("absence" NEXT seizure*):ti,ab OR "seizure absence":ti,ab OR ("complex partial" NEXT seizure*):ti,ab OR "partial seizure complex":ti,ab OR "partial seizures complex":ti,ab OR "seizure complex partial":ti,ab OR ("atonic" NEXT seizure*):ti,ab OR "seizure atonic":ti,ab | 9,428 |
| 3. | #1 AND #2 | 26 |

### Table S2- Quality assessment of the cohorts using the Newcastle-Ottawa scale (NOS) modified for cohort studies.

| **Study** | **Selection (Maximum 4 stars)** | | | | **Comparability (Maximum 2 stars)** | **Outcome (Maximum 3 stars)** | | | Total scores |
|---|---|---|---|---|---|---|---|---|---|
| | Representativeness of the exposed cohort | Selection of the nonexposed cohort | Ascertainment of exposure | Demonstration that outcome of interest was not present at start of study | Comparability of cohorts on the basis of the design or analysis | Assessment of outcome | Was follow-up long enough for outcomes to occur | Adequacy for follow-up of cohorts | |
| Romozzi et al. | * | * | * | | * | * | * | * | 7 |
| Martinez-Fernandez et al. | * | | * | * | | * | * | * | 6 |
| Núñez et al. | * | | * | * | | * | * | * | 6 |
| Fang et al. | * | | * | * | | | * | * | 6 |
| Yang et al. | * | * | | * | * | | * | * | 6 |
| Lu Q et al. | * | | * | * | | * | * | * | 6 |
| Huang et al | * | | * | * | | * | * | * | 6 |
| Pang et al. | * | | * | * | | * | * | * | 6 |
| Wang et al. | * | * | * | * | ** | * | * | * | 9 |

### Table S3- Quality assessment of the cross-sectional studies using the Newcastle-Ottawa scale (NOS) modified for cross-sectional studies.

| Study | Selection: (Maximum 5 stars) | | | | Comparability: (Maximum 2 stars) | Outcome: (Maximum 3 stars) | | |
|---|---|---|---|---|---|---|---|---|
| | Representativeness of the sample | Sample size | Non-respondents | Ascertainment of the exposure (risk factor): | The subjects in different outcome groups are comparable, based on the study design or analysis. Confounding factors are controlled | Assessment of the outcome | Statistical test | Total scores |
| Li et al. | * | * | | * | ** | * | * | 7 |
| Massoud et al. | * | * | | ** | ** | * | * | 8 |
| Lu L et al. | * | * | | ** | ** | * | * | 8 |
| von Wrede et al. | * | * | | * | ** | * | * | 7 |
| Kadali et al. | * | * | | ** | ** | * | * | 8 |
| Chan et al. | * | * | * | ** | * | ** | | 8 |
| Özdemir et al. | * | * | * | ** | ** | * | * | 9 |
| Hood et al. | * | * | * | * | ** | * | * | 8 |
| King et al. | | | | * | ** | * | | 4 |

### Figure S1- PRISMA flow diagram of the systematic search

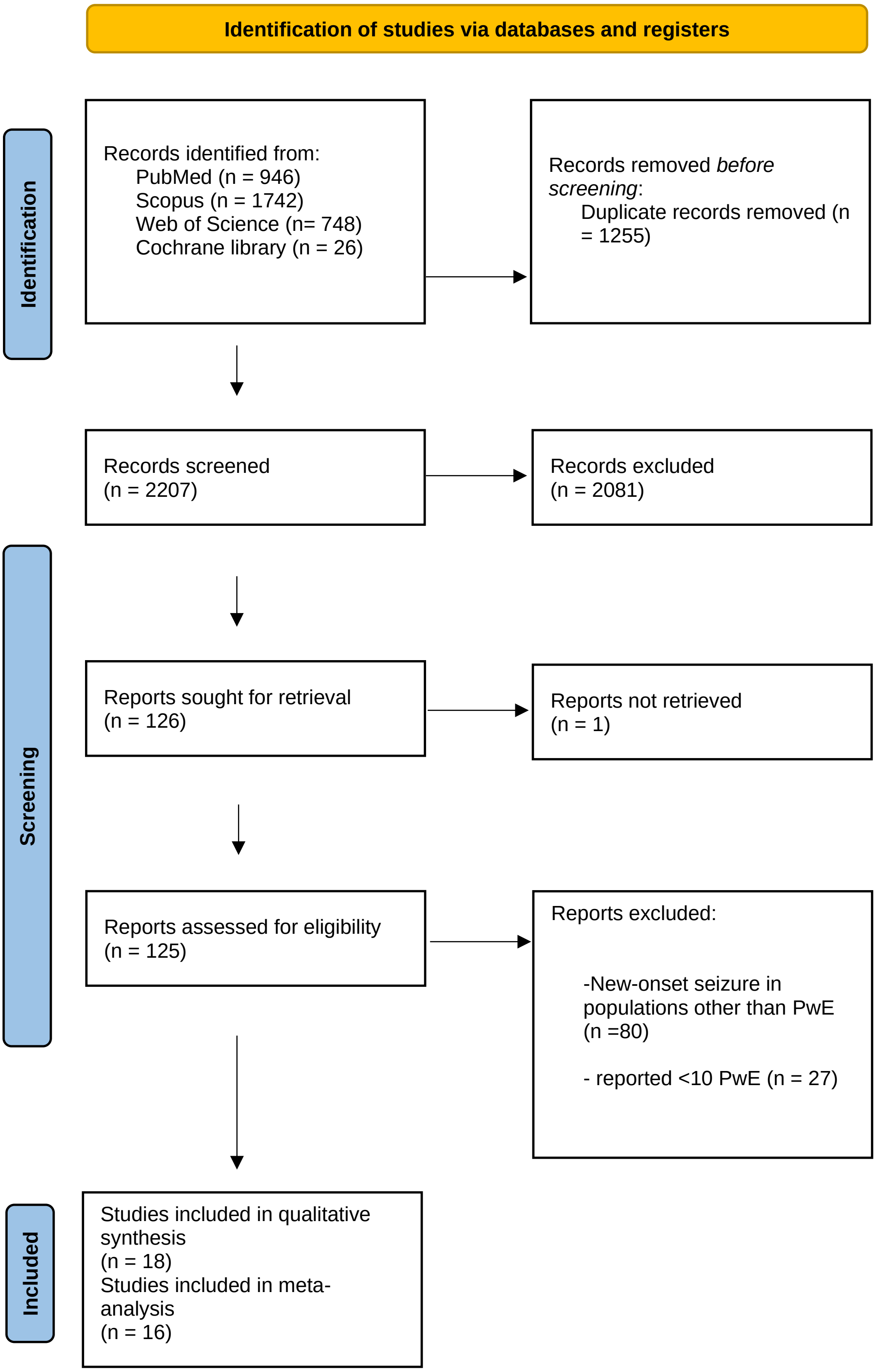

### Figure S2- Funnel plot for studies reporting increased seizure frequency after COVID-19 vaccines.

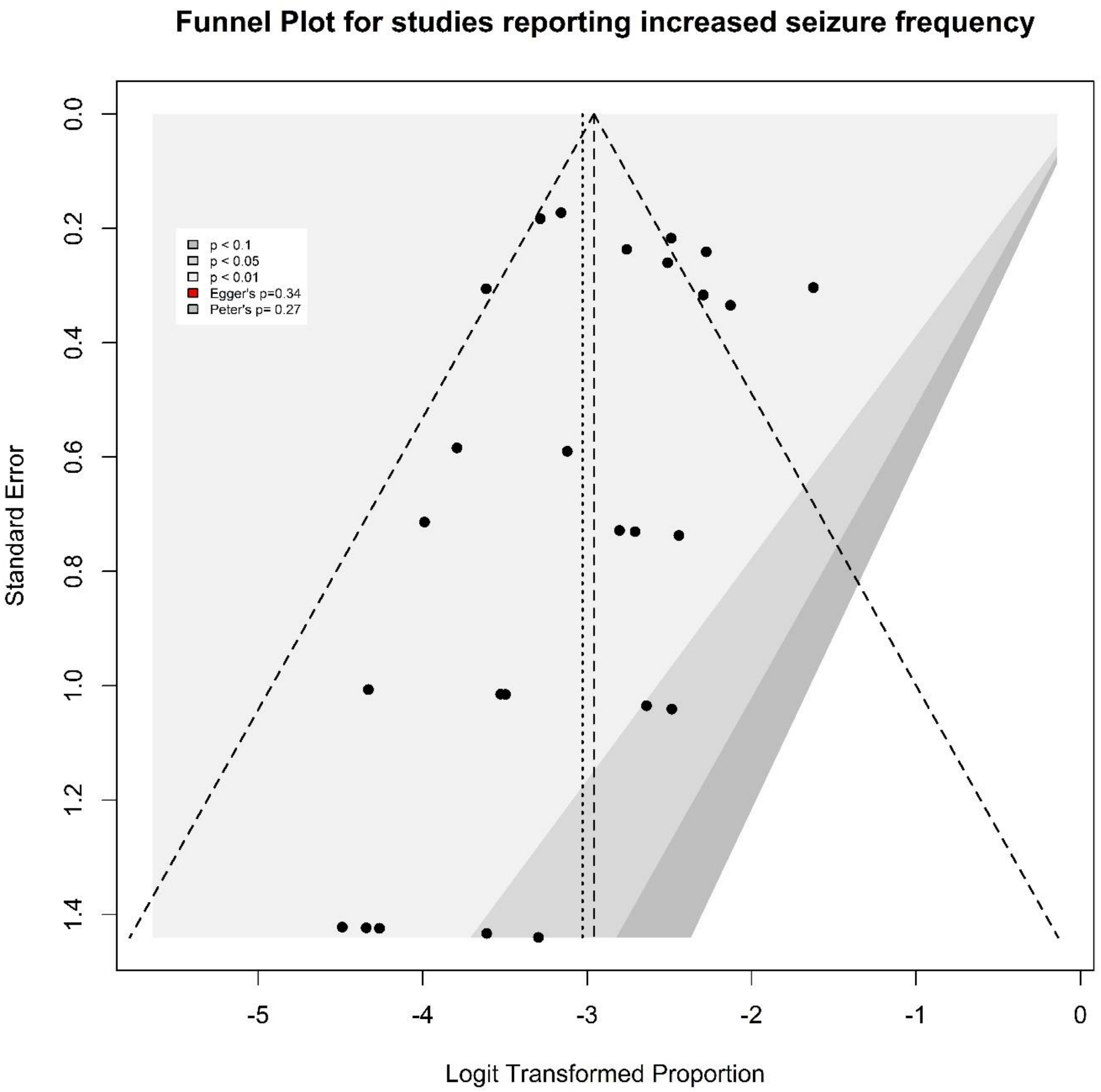

### Figure S3- Funnel plot for studies comparing mRNA vs. viral-vector COVID-19 vaccines regarding increased seizure frequency.

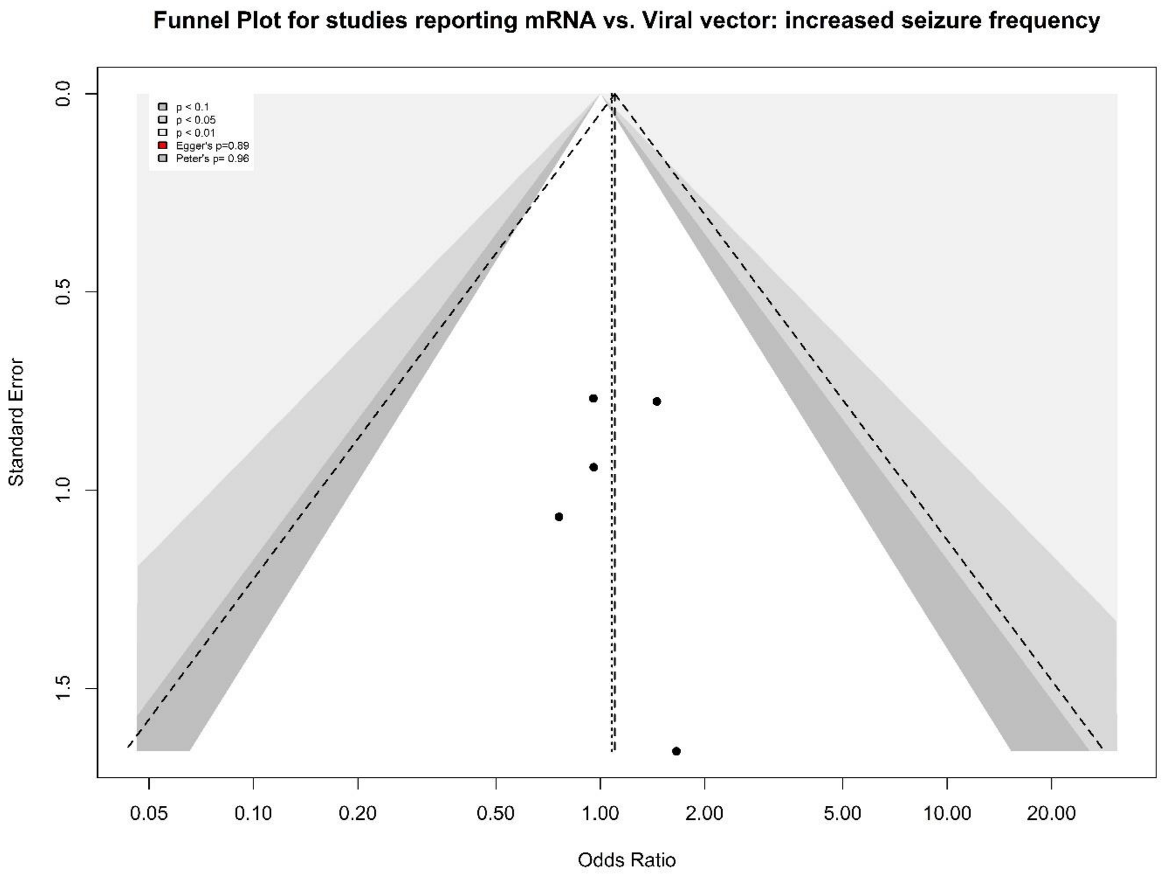

**Figure S4- Funnel plot for studies reporting status epilepticus incidence proportion after COVID-19 vaccines.**

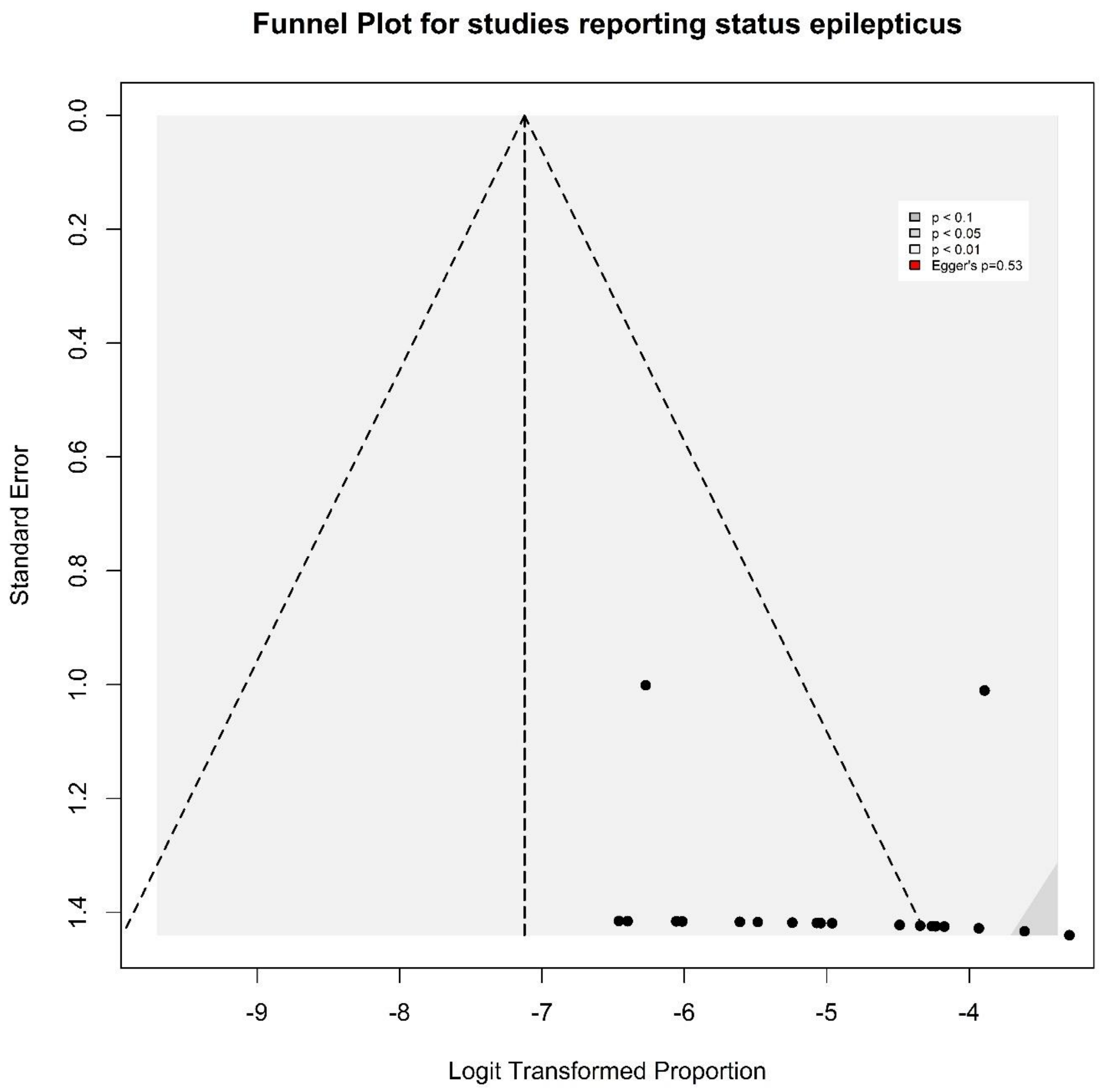
